# Development and Validation of a Deep Learning System for the Diagnosis of Pediatric Diseases: A Large-Scale Real-World Data Study in Shanghai

**DOI:** 10.1101/2022.10.07.22280541

**Authors:** Xiaoling Ge, Yi Wang, Li Xie, Yujuan Shang, Yihui Zhai, Zhiheng Huang, Jianfeng Huang, Chengjie Ye, Ao Ma, Wanting Li, Xiaobo Zhang, Hong Xu

## Abstract

**Background:** Artificial intelligence (AI)-assisted diagnosis is considered to be the future direction of improving the efficiency and accuracy of pediatric diseases diagnosis, while the existing research based on AI are far from sufficient because of limited data amount, inadequate coverage of disease types, or high construction costs, and have not been applied on a large scale. We aimed to develop an accurate deep learning model trained on millions of real-world data to verify the feasibility of the technology, and build the whole process of outpatient auxiliary diagnosis.

**Methods and findings:** We applied a Chinese Natural Language Processing (NLP) and an end-to-end deep neural network classifier to the outpatient’s electronic medical records (EMRs) in a single child care center in Shanghai, China, to unstructured text processing and construct an auxiliary diagnostic model, all patients were aged from 0 to 18 years. A training cohort with millions of records and an independent validation cohort with tens of thousands of records were intake separately and calculate diagnosis concordance rate (DCR) of model in each diseases group. The records with inconsistent diagnoses between human and AI were evaluated by clinical experts’ group, and calculate the relative correct rate (RCR) to evaluate the diagnostic performance of the model. A total of 5,271,347 medical records were intake in model training covering sixteen categories of diseases according to disease coding, reaching a DCR of 95· 49% (95· 48∼95· 51). For validation, 91,880 records were obtained from validation dataset, which reached a DCR of 93· 51% (93· 35∼93· 67) and FDCR of 72.04% (71· 75∼72· 33). It was confirmed that the accuracy of the model was still higher than that of human with most RCR>1 in validation dataset.

**Conclusions:** The deep learning system could support diagnosis of pediatric diseases, which has high diagnostic performance, comprehensive disease coverage, feasible technology, and can be promoted in multiple sites in the future.

**Funding:** The Authors received no specific funding for this work.

## Introduction

Diagnoses that are missed, incorrect or delayed, which are often less mentioned than problems such as drug errors and operational mistakes, are believed to affect 10%-20% of cases and result in serious consequences. The expression ability of pediatric patients is poor, and the clinical manifestations of diseases are atypical. In addition, physicians specializing in adult medicine are frequently requested to play the role of a pediatrician in grassroots hospitals, even though they tend to lack of knowledge and experience in pediatrics, and have subjective assumptions and narrow ideas in clinical thinking.^1^ Unfortunately, most clinicians tend to avoid discussing diagnostic errors publicly, which makes it difficult to detect and correct them promptly.^2^

As we learn more about pediatric diseases, we learn more about the power of artificial intelligence (AI) tools that can be used in unprecedented ways. AI-assisted diagnosis is considered to be an important means to summarize complex diagnostic mechanisms, improve the efficiency and accuracy of pediatric clinical diagnostics,^3^ and provide a solution to the imbalance between the supply and demand of pediatricians and patients sequentially. The deep learning method, as a subgroup of AI, is a particularly promising method that automatically learns entity representations from natural language and has been shown to match and even outperform human performance in task-specific applications. Although it requires large datasets for training, deep learning has demonstrated relative robustness to noise in ground truth labels, among others. The automated capabilities of AI offer the potential to enhance the qualitative expertise of clinicians, including improving diagnostic accuracy.

Many clinically intelligent decision support products, such as HM Healthcare, iFLYTEK Co.Ltd intelligent medical assistant, and Baidu 01 Healthcare, have emerged and been applied in many primary hospitals in recent years in China. However, these studies mainly focus on adult diseases and lack learning at the pediatric level. In the field of pediatrics, AI technology has been applied to research on special disease auxiliary diagnosis of children’s respiratory diseases,^4^ sepsis, and noninfectious systemic inflammatory response syndrome (SIRS)^5^ in foreign countries; however, there is no research on general pediatric auxiliary diagnosis and treatment yet. In 2019, a deep learning framework that analyzes more than 140,000 electronic medical records (EMRs) to study intelligent diagnosis, including the diagnosis of 63 pediatric diseases, using Chinese EMRs was proposed by Wu et al.^6,7^ Moreover, Liang et al.^8^ developed an AI intelligent diagnosis model that learned 1· 36 million high-quality electronic text medical records and constructed an AI-assisted diagnosis model that has an accuracy of nearly 90% in the diagnosis of 55 common pediatric diseases. However, the existing diagnostic models based on AI are far from sufficient in terms of learning data or diseases categories, and are still limited in the diagnosis of children’s diseases and have not been applied on a large scale.

To solve the pain points of the intelligent decision-making research above, the EMRs of millions of outpatients in a single-central hospital for children in Shanghai, China, Chinese Natural Language Processing (NLP) and an end-to-end deep neural network classifier were used for unstructured text processing and learning to imitate the deductive reasoning process of the doctor’s brain’s assumptions. A deep learning system for pediatric diseases based on AI was developed for initial diagnosis in the pediatric outpatient department in this study.

## Methods

### Training Dataset of the Deep Learning System (DLS)

The study began in March 2021. All outpatient visits records of patients up to eighteen years of age between Jan 1, 2017, and Aug 10, 2020 in the Children’s Hospital of Fudan University, were included in this study for model training, regardless of sex, disease group, or disease progression. Department of visit, month of visit, sex, age, chief complaint, physical examination, disease history, and primary diagnosis were extracted from the EMR database of the Big Data Management System. This study was approved by the Research Ethics Committee of the Children’s Hospital of Fudan University. Informed consent was waived for retrospectively collected outpatient medical record data, which were anonymized. None of the researchers were able to identify individual participants during or after data collection.

### Data Cleaning

We excluded records meeting the following criteria (Figure 1): 1) records with missing/blank fields; 2) records with duplicated information due to template use; and 3) records with information less than twenty words. We obtained 5,271,347 patient visit records for model training after filtering. These records cover 306 fine-grained medical majors in the hospital within the time frame of inclusion.

**Fig 1.**
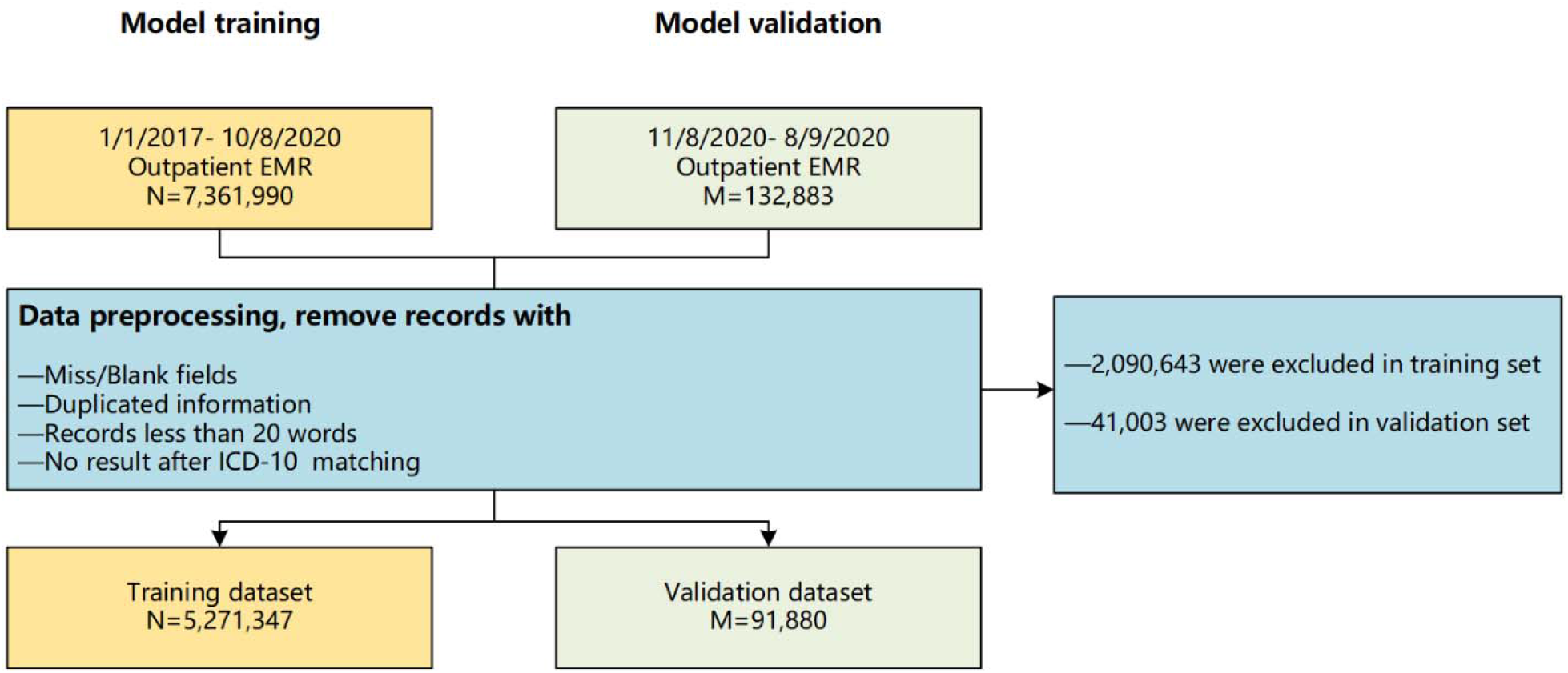
Flow chart of study population inclusion.

### Data Labeling

The codes from the International Statistical Classification of Diseases and Related Health Problems, 10^th^ Revision (ICD-10) were assigned to each record by exact word matching of doctor diagnosis in Chinese. Multiple diagnoses were allowed in which case the first diagnosis was considered the most important main diagnosis. All cases were divided into 16 categories of diseases based on ICD-10 coding categories, and the codes derived from pregnancy, childbirth, puerperium (O00-O99), or other diseases without obvious anatomical classification (R00-Z99) were classified as “other diseases”.

### NLP Model Construction

First, department of visit, age, sex, chief complaint, physical examination, and disease history are concatenated into one sentence as input. Our NLP DLS consists of two parts: the Chinese language feature extractor and the end-to-end deep neural network classifier. The Chinese language feature extractor extracts 1–4 n-grams of Chinese characters as well as possible number/alphabet characters. For a sentence of length T, there will be 4T-6 n-grams as features. We embed this one-hot n-gram into a hidden space of 512 dimensions. The embedding matrix is initialized with public word vectors from Tencent.^9^ For unknown n-grams, a random initialization of N (0,0· 1) is applied.

The end-to-end deep neural network consists of three fully connected hidden layers, each of which consists of 512 hidden units. A softmax layer is employed as a loss layer. A special modification is applied on the standard softmax layer to allow multiple diagnosis labels. For m diagnoses, we arbitrarily weight each diagnosis as m, m-1, m-2…1 because we consider the first diagnosis as the main and most important diagnosis. We then normalize the weights to 1 and feed them into softmax as “soft labels”.

Stochastic gradient decent (SGD) is employed to train the neural network. We perform input dropout at a rate of 50% to regularize the network. However, we turn off dropout in the hidden layers. During the training, 5% of the training dataset is held out to monitor the training process. The training is stopped when the loss on held-out samples stops improving. All the frameworks were coded by the author from scratch using the C++ programming language. When the DLS is connected to the EMR system, after the doctor fills in chief complaint, physical examination, and disease history, the model will quickly complete the calculation and provide the doctor with diagnosis opinion in the form of ‘probability value, ICD-10 code, diagnosis name’ list. The construction process of deep learning system is shown in Figure 2.

**Fig 2.**
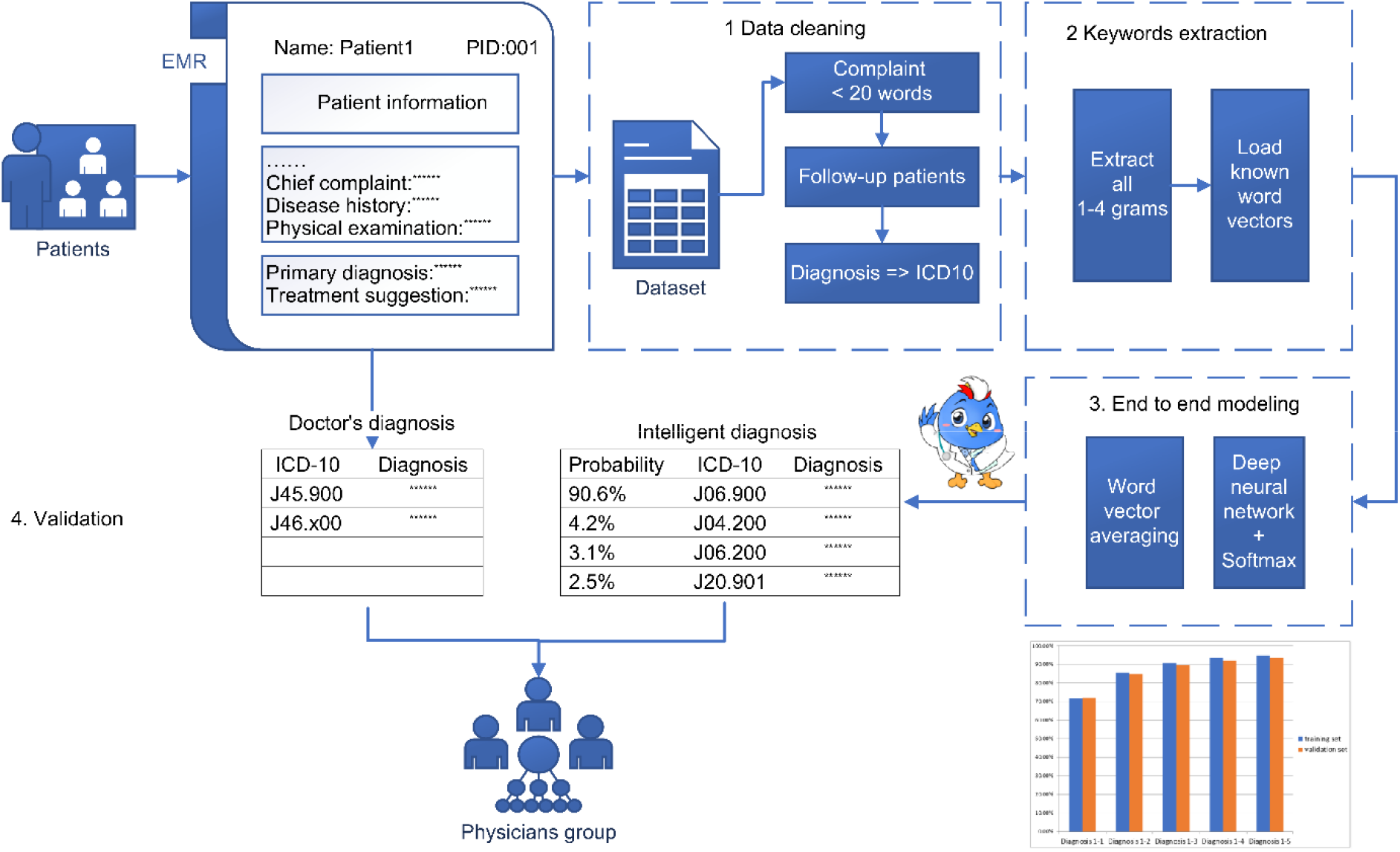
Workflow of the deep learning algorithm. Department of visit, age, sex, chief complaint, physical examination, and disease history are concatenated into one sentence as input dataset. (1) Excluded records meeting the following criteria: a) records with missing/blank fields; b) records with duplicated information due to template use; and c) records with information less than twenty words. (2) The Chinese language feature extractor extracts 1–4 n-grams of Chinese characters as well as possible number/alphabet characters. (3) The end-to-end deep neural network. The embedding matrix is initialized with public word vectors from Tencent. (4) In the clinical evaluation, an expert group was used to evaluate the parts with great differences between human and machine diagnosis results collectively.

### Validation Dataset

All outpatient visit records of patients up to eighteen years of age between August 11, 2020 and September 8, 2020 in Children’s Hospital of Fudan University, were included in this study for model testing, regardless of sex, disease group, or disease progression. Department of visit, month of visit, sex, age, chief complaint, physical examination, disease history, and primary diagnosis were extracted from the Big Data Management System database (Validation dataset). After the same filtering pipeline as the training dataset, 91,880 records were obtained for testing.

### Comparison of the Performance of AI System with Human Physicians

Referring to the man-machine diagnosis comparison of the Dxplain clinical decision support system (CDSS), an expert group composed of one senior expert with more than five years of experience and one expert at or above the associate chief physician level with more than ten years of experience, was invited to evaluate the parts with great differences between human and machine diagnosis results collectively in each clinical department during the clinical evaluation phase. Each encountered medical record was assigned the same number of AI diagnoses based on the number of diagnoses given by the doctor. Experts may choose the more accurate diagnosis by blind selection between the human diagnosis and the AI diagnosis based on medical record information consisting of department of visit, month of visit, sex, age, chief complaint, physical examination, and disease history. There are four possible outcomes in clinical evaluation: (1) AI Correct: AI’s diagnosis is better than that of the doctor’s; (2) Physician Correct: the doctors’ diagnosis is better than that of AI; (3) Both Correct: both diagnoses are correct; and (4) Invalid: both diagnoses are incorrect.

### Statistical Analysis

We defined the following three measurements of AI diagnostic accuracy.

1) First diagnostic concordant rate (FDCR): Top-1 accuracy, the percentage of records in the testing dataset in which any of the AI’s top 1 diagnoses are contained in the human doctors’ diagnoses.

2) Diagnosis concordance rate (DCR): Top-5 accuracy, the percentage of records in the testing dataset in which any of the AI’s top 5 diagnoses are contained in the human doctors’ diagnoses.

3) Relative correct rate (RCR): In a blind competition between AI and human doctors, The RCR is defined as:

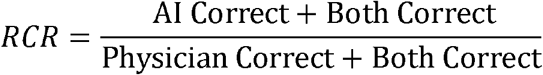

When RCR>1, the diagnostic ability of AI is stronger than that of the doctor, and when RCR<1, the diagnostic ability of AI is weaker than that of the doctor.

## Results

### Baseline Information

In the training dataset, a total of 7,361,990 outpatient EMRs were collected in the outpatient EMR system from the Children’s Hospital of Fudan University. After processing, 5,271,347 medical records were included in model training, covering over 300 kinds of outpatient special clinics. The median age was approximately 4·58 years (0–18 years), and 56·09% of the patients were male, which was a higher percentage than female patients. Among the sixteen categories of diseases, respiratory diseases were the most common (34·53%), followed by infectious diseases (9·34%) and skin-related diseases (6·80%).

To validate the model, 91,880 records were obtained from EMR. There were generally more boys than girls, which was the same as the sex distribution in the training set. As shown in Table 1, there were obvious differences in the composition of disease types between the test dataset and the training set, but respiratory diseases were also dominant.

**Table 1.**
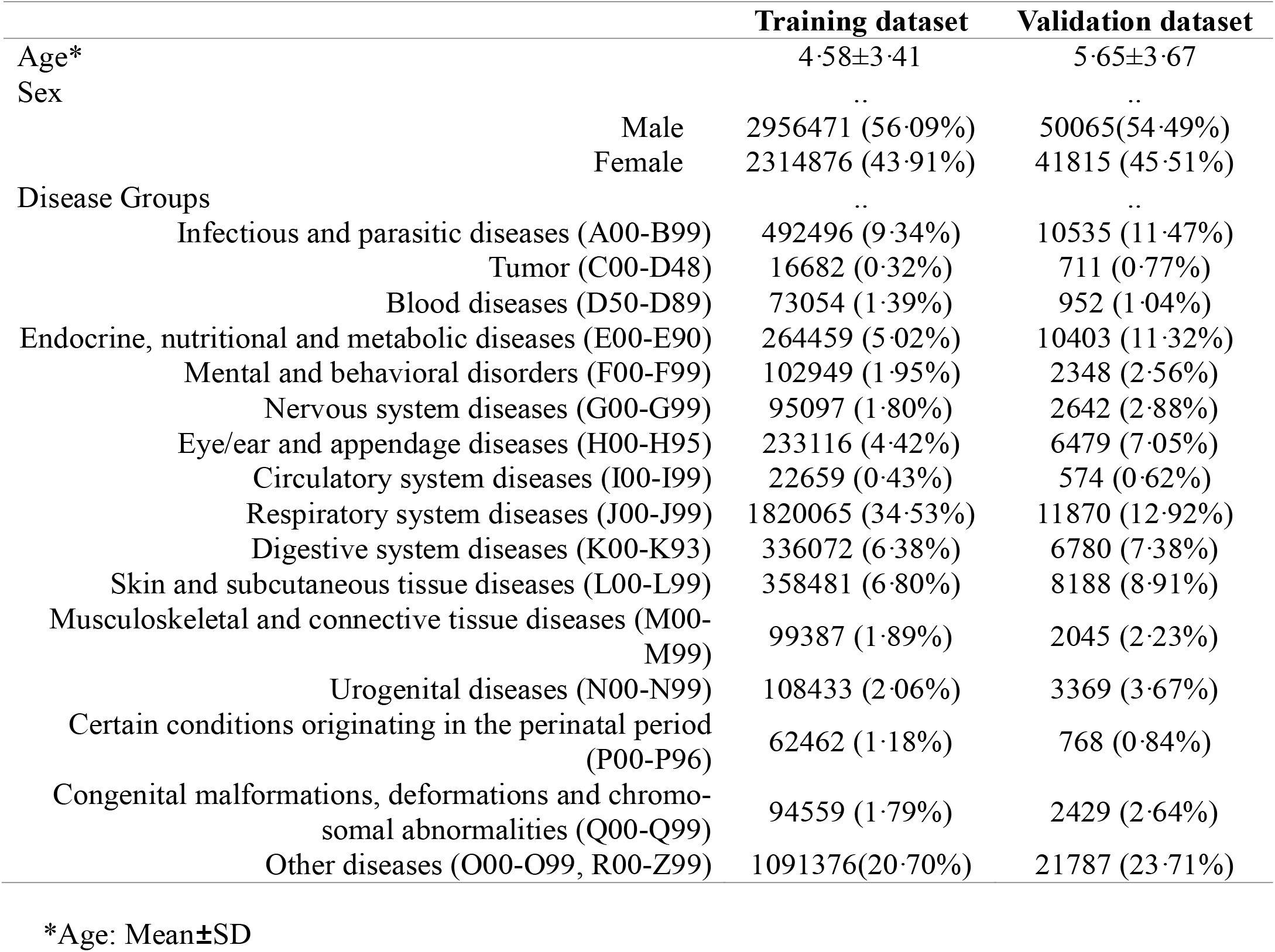
Basic Data of Outpatient Electronic Medical Records.

### Performance of the AI Models

In the experiment, we focused on the diagnostic performance of the AI model in each type of disease; that is, to explore the consistency rate of the model with the existing doctor’s diagnosis in different diseases, after model training, the performance of the model in different diseases was evaluated by calculating the DCR in the two datasets (Table 2), and the corresponding 95% confidence interval was calculated. Subsequently, for cases where the AI and the doctors disagreed, a third-party judgment was made by a clinical expert group in each specialty to explore who had a more accurate diagnosis.

**Table 2.** Illustration of the Diagnostic Performance of the AI Model (training dataset)

| Diseases groups | N | DCR % | 95% CI |
| --- | --- | --- | --- |
| Infectious and parasitic diseases (A00-B99) | 492496 | 96.70 | (96.65~96.75) |
| Tumor (C00-D48) | 16682 | 75.21 | (74.56~75.87) |
| Blood diseases (D50-D89) | 73054 | 92.81 | (92.62~93.00) |
| Endocrine, nutritional and metabolic diseases (E00-E90) | 264459 | 97.11 | (97.04~97.17) |
| Mental and behavioral disorders (F00-F99) | 102949 | 94.70 | (94.56~94.83) |
| Nervous system diseases (G00-G99) | 95097 | 92.55 | (92.38~92.72) |
| Eye/ear and appendage diseases (H00-H95) | 233116 | 98.26 | (98.21~98.31) |
| Circulatory system diseases (I00-I99) | 22659 | 80.13 | (79.62~80.65) |
| Respiratory system diseases (J00-J99) | 1820065 | 96.86 | (96.84~96.89) |
| Digestive system diseases (K00-K93) | 336072 | 94.31 | (94.23~94.39) |
| Skin and subcutaneous tissue diseases (L00-L99) | 358481 | 95.36 | (95.29~95.43) |
| Musculoskeletal and connective tissue diseases (M00-M99) | 99387 | 87.62 | (87.42~87.83) |
| Urogenital diseases (N00-N99) | 108433 | 93.16 | (93.01~93.31) |
| Certain conditions originating in the perinatal period (P00-P96) | 62462 | 93.05 | (92.85~93.25) |
| Congenital malformations, deformations and chromosomal abnormalities (Q00-Q99) | 94559 | 88.84 | (88.64~89.05) |
| Other diseases (O00-O99, R00-Z99) | 1091376 | 94.90 | (94.86~94.94) |
| All diseases | 5271347 | 95.49 | (95.48~95.51) |

DCR%: Diagnosis concordance rate, the percentage of records in the testing dataset in which any of the AI’s top 5 diagnoses are contained in the human doctors’ diagnoses. 95%CI: 95% confidence interval.

#### Diseases groups N DCR % 95% CI

After over five million cases of training, the overall diagnostic consistency rate reached 95·49% (95·48∼95·51). Among the different diseases, the models performed best in eye/ear and appendage diseases (H00-H95). On the other hand, the DCRs were relatively low in diagnoses of categories such as tumor (C00-D48), circulatory system diseases (I00-I99), musculoskeletal and connective tissue diseases (M00-M99), and any other congenital diseases (Q00-Q99), with DCRs of less than 90%.

### Model Validation

In local validation, the overall DCR reached 93·51% (93·35∼93·67) while FDCR was 72.04% (71·75∼72·33), and the AI model performed better in eye/ear and appendage diseases(H00-H95), infectious and parasitic diseases (A00-B99), endocrine, nutritional and metabolic diseases (E00-E90), and respiratory system diseases (J00-J99), with DCRs exceeding 95%. In contrast, it performed poorly in tumor (C00-D48) and circulatory system diseases (I00-I99), with DCRs lower than 80%. In addition, the model did not perform well in the first diagnosis of Circulatory system diseases (I00-I99) and Certain conditions originating in the perinatal period (P00-P96), but the accuracy was significantly improved after expanded to the top five diagnoses. There were significant differences in the performance of the model in the training set and validation set even if the cases came from the same hospital, especially in the tumor, blood diseases, and certain conditions originating in the perinatal period (>5%). In general, the DCRs of most disease types in the test set were lower than those of the training set, but the DCRs of some disease types were slightly higher, such as in infectious and parasitic diseases (A00-B99), as well in eye/ear and appendage diseases (H00-H95).

### Third-party Evaluation

The medical records with inconsistent diagnoses between the AI model and the doctor were evaluated by clinical experts. According to the evaluation principle, two doctors in each specialty jointly evaluated the cases and provided the evaluation results of Validation set. Due to the differences in disease distributions and AI diagnostic capabilities in different diseases, the number of cases in the validation set was inconsistent. Based on the results of the relative accuracy calculation (Table 3), experts believe that the accuracy of the model was generally higher than that of doctors.

**Table 3.** Illustration of the Diagnostic Performance of the AI Model (validation dataset)

| Diseases groups | N | FDCR % | 95% CI | DCR % | 95% CI |
| --- | --- | --- | --- | --- | --- |
| Infectious and parasitic diseases (A00-B99) | 10534 | 81.69 | (80.95~82.43 ) | 97.07 | (96.74~97.39) |
| Tumor (C00-D48) | 707 | 38.05 | (34.47~41.63) | 68.46 | (65.03~71.89) |
| Blood diseases (D50-D89) | 952 | 67.54 | (64.57~70.52) | 85.29 | (83.04~87.54) |
| Endocrine, nutritional and metabolic diseases (E00-E90) | 10403 | 71.75 | (70.88~72.61) | 96.68 | (96.34~97.03) |
| Mental and behavioral disorders (F00-F99) | 2346 | 67.22 | (65.32~69.12) | 90.41 | (89.22~91.60) |
| Nervous system diseases (G00-G99) | 2642 | 79.75 | (78.22~81.28) | 91.68 | (90.62~92.73) |
| Eye/ear and appendage diseases (H00-H95) | 6478 | 87.09 | (86.28~87.91) | 98.32 | (98.00~98.63) |
| Circulatory system diseases (I00-I99) | 574 | 44.43 | (40.36~48.49) | 76.13 | (72.65~79.62) |
| Respiratory system diseases (J00-J99) | 11869 | 69.21 | (68.38~70.04) | 95.69 | (95.32~96.05) |
| Digestive system diseases (K00-K93) | 6777 | 73.60 | (72.55~74.65) | 93.43 | (92.84~94.02) |
| Skin and subcutaneous tissue diseases (L00-L99) | 8188 | 68.45 | (67.45~69.46) | 93.61 | (93.08~94.14) |
| Musculoskeletal and connective tissue diseases (M00-M99) | 2038 | 63.69 | (61.60~65.78) | 83.37 | (81.75~84.98) |
| Urogenital diseases (N00-N99) | 3369 | 74.56 | (73.09~76.03) | 92.52 | (91.63~93.41) |
| Certain conditions originating in the perinatal period (P00-P96) | 768 | 55.99 | (52.48~59.50) | 86.20 | (83.76~88.64) |
| Congenital malformations, deformations and chromosomal abnormalities (Q00-Q99) | 2426 | 65.62 | (63.73~67.51) | 84.79 | (83.36~86.22) |
| Other diseases (O00-O99, R00-Z99) | 21781 | 68.73 | (68.11~69.35) | 92.14 | (91.79~92.50) |
| All diseases | 91852 | 72.04 | (71.75~72.33) | 93.51 | (93.35~93.67) |

FDCR: First diagnosis concordance rate, the percentage of records in the testing dataset in which any of the AI's top 1 diagnoses are contained in the human doctors' diagnoses. DCR: Diagnosis concordance rate, the percentage of records in the testing dataset in which any of AI's top 5 diagnoses are contained in the human doctors' diagnoses. 95% CI: 95% confidence interval.

**Table 3.** Validation of the Diagnostic Performance of the AI Model and Physicians.

| Diseases groups | Model validation at the research site |  |  |  |  |  |
| --- | --- | --- | --- | --- | --- | --- |
|  | N | AI Cor-<br>rect | Physician<br>Correct | Both<br>Correct | Invalid | RCR |
| Infectious and parasitic diseases (A00-B99) | 82 | 25 | 15 | 18 | 24 | 1.30 |
| Tumor (C00-D48) | 31 | 9 | 5 | 4 | 13 | 1.44 |
| Blood diseases (D50-D89) | 21 | 9 | 3 | 0 | 9 | 3.00 |
| Endocrine, nutritional and metabolic diseases (E00-E90) | 57 | 19 | 16 | 6 | 16 | 1.14 |
| Mental and behavioral disorders (F00-F99) | 27 | 12 | 3 | 4 | 8 | 2.29 |
| Nervous system diseases (G00-G99) | 43 | 13 | 9 | 3 | 18 | 1.33 |
| Eye/ear and appendage diseases (H00-H95) | 43 | 12 | 12 | 6 | 13 | 1.00 |
| Circulatory system diseases (I00-I99) | 20 | 5 | 1 | 1 | 13 | 3.00 |
| Respiratory system diseases (J00-J99) | 86 | 45 | 11 | 10 | 20 | 2.62 |
| Digestive system diseases (K00-K93) | 87 | 28 | 18 | 14 | 27 | 1.31 |
| Skin and subcutaneous tissue diseases (L00-L99) | 110 | 33 | 25 | 7 | 45 | 1.25 |
| Musculoskeletal and connective tissue diseases (M00-M99) | 22 | 7 | 4 | 6 | 5 | 1.30 |
| Urogenital diseases (N00-N99) | 48 | 12 | 10 | 3 | 23 | 1.15 |
| Certain conditions originating in the perinatal period (P00-P96) | 20 | 8 | 0 | 4 | 8 | 3.00 |
| Congenital malformations, deformations and chromosomal abnormalities (Q00-Q99) | 73 | 21 | 11 | 13 | 28 | 1.42 |
| Other diseases (O00-O99, R00-Z99) | 290 | 94 | 65 | 37 | 94 | 1.28 |

AI: artificial intelligence, RCR: Relative correct rate = (AI Correct + Both Correct)/(Physician Correct + Both Correct). When RCR>1, the diagnostic ability of AI is stronger than that of the doctor, and when RCR<1, the diagnostic ability of AI is weaker than that of the doctor.

## Discussion

In this study, we developed and validated a pediatric diagnosis CDSS based on a deep learning system. Utilizing electronic health records, AI achieved high performance with satisfying DCRs and RCR. In the prospective validation dataset, AI outperformed physicians from a tertiary hospital. In these two datasets, there were generally more boys than girls, which is the same as the distribution in other similar studies.^8^ There were obvious differences in the mean age and composition of disease types among the two datasets, which may be because the validation dataset was taken from a continuous month, resulting in a bias caused by seasonal factors.

Although FDCR (top-1 accuracy) is only 72.04% (71· 75∼72· 33), DCR (top-5 accuracy) has a significant improvement of 93· 51% (93· 35∼93· 67). Considering the complexity of the disease and the compatibility of diagnostic codes, there are more than one diagnostic choice in a case as a multi-label classification problem. This study mainly focused on the DCR index. As shown in the results, the AI model performed better in internal medical diseases, including infectious and parasitic diseases, endocrine/nutritional and metabolic diseases, eye/ear and appendage diseases, respiratory system diseases, etc., and best performance was in eye/ear and appendage diseases, which was because of the more concentrated distribution of common diseases in the clinic. However, in surgical diseases, such as musculoskeletal and connective tissue diseases, the DCR was lower. This can be explained because surgical diseases often need to be combined with various laboratory tests and radiographic examinations for auxiliary diagnosis in the clinic. Therefore, AI diagnosis was relatively correct and could provide a reference for local doctors. Pediatric medical resources are scarce and unevenly distributed in China. As of 2015, the shortage of pediatricians was as high as 200,000, and the ratio of pediatric doctors to patients was 1:3500 in China, which is far lower than the 1:1000 doctor-patient standard in developed countries.^10^ Similar situations are common in Europe,^11^ the US^12^ and Japan.^13^ In addition, the excessive concentration of high-quality medical resources leads to the exposure of primary pediatricians to fewer diseases and cases, and the lack of clinical experience results in a high incidence of misdiagnoses and diagnostic errors in primary hospitals, which delays timely access to treatment for patients. In 2013, a survey report from the American Academy of Pediatrics’ Quality Improvement Innovation Networks (QuIIN) showed that 35% of pediatricians may make a diagnostic error at least once a month, and 33% have a diagnostic error at least once a year, finally resulting in adverse events.^14^ It is supposed that the insufficient clinical knowledge of doctors, the incorrect collection of consultation information, and invalid inspection and verification are the main reasons for clinical misdiagnosis.^15^ However, when retrieving the relevant research at home and abroad in the last ten years based on the keywords “Patient safety” and “Diagnostic Errors” in PubMed, only 6% focused on the diagnostic safety of children. A previous study reported that the accuracy rate of the first diagnosis of pediatricians in the primary hospital was relatively low.^16^ In addition, the diagnostic accuracy of primary units at lower professional levels (e.g., level 1 and level 2) was lower than that of level 3. Research on reducing the pediatric misdiagnosis rate is far behind the progress of adult and other patient safety fields.^17^ Regarding the abovementioned status, many suggested strategies to reduce the incidence of diagnostic errors have been proposed, including improving the clinical expertise of clinicians and reducing the inherent cognitive errors of doctors to make better decisions.^15^ Therefore, the application of AI systems could be beneficial among areas where healthcare providers are in a relative shortage. It will greatly increase the accuracy of the first diagnosis and thus significantly improve medical health care in Chinese children. In addition, this AI model could provide a new idea for the quality control of EMRs in the outpatient departments of local hospitals.

The AI system demonstrates high diagnostic accuracy across multiple diseases and is comparable to well-trained pediatricians in diagnosing common pediatric diseases. In the past, the CDSS was widely used to extract key clinical information based on EMRs for reasoning, which could help clinicians assess disease status, make diagnoses, choose appropriate treatments and make other clinical decisions.^18^ The clinical diagnosis and treatment guidelines and a large number of literary studies have provided a reliable source of knowledge for the system and have been universally recognized. However, there are few systems that have been put into extensive and long-term application in clinical practice worldwide. On the one hand, the construction of a knowledge base cannot meet the needs of clinicians; furthermore, most systems are not technically integrated with EMRs, resulting in disconnection from the clinical workflow, which reduces the enthusiasm of clinicians to use the CDSS. With the rapid development of medical information technology, AI technology has become a powerful support for the healthcare revolution. With patients as the center and medical institutions as the main service body, medical AI can cover the whole process from disease prevention, diagnosis, and treatment to patient rehabilitation by using AI technologies such as knowledge mapping or deep learning.^19^ At the same time, with the widespread application of EMRs in recent years, combined with large data-level EMRs and other related information learning, AI algorithms can complete complex analysis tasks in a short period of time, feedback the best classification model results based on the input information, and assist doctors in improving the accuracy and efficiency of patient diagnosis. “Data-driven” intelligent aid decision-making based on real-world EMR data will have the potential to supplement traditional rule-based decision-making methods.^20^

## Limitations

First, this AI system is only suitable for identifying the diagnosis at first clinic at the present stage. Second, results of laboratory medicine and imaging examination were not included in the AI system. Third, this AI system cannot provide disease severity classification or treatment suggestions yet. Fourth, this study lacks a comparison of multiple model algorithms.

## Conclusions

In our study, we developed and validated an AI-based system that can provide clinical decision support in the event of diagnostic uncertainty and complexity. The AI model can quickly and accurately identify the diagnosis of children, which can help pediatricians make more precise diagnoses while further preventing undetected cases. Our findings are of great clinical value and practical significance in improving the health care of children in China and optimizing medical resources.

## Data Availability

The institutional data used for training and validation are not publicly available, because they contain protected patient health information. Source code of the deep neural network can be made available, subject to intellectual property constraints, by contacting the co-first author.

## Contributors

XG, YW, XZ, HX conceived of and designed the study. XG, YW, YS, AM, CY, WL, YZ, ZH, JH, XZ and H X contributed to Acquisition, analysis, or interpretation of data. XG, YW, LX, YS, XZ, HX contributed to draft of the manuscript, which was approved by all the authors. XG, YW, YS, YZ, XZ, HX contributed to critical revision of the manuscript for important intellectual content. XG, YW, LX, YS contributed to statistical analysis. XZ, HX contributed to supervision. HX and XZ had full access to all the data in the study and take responsibility for the integrity of the data and the accuracy of the data analysis. XG and YW contributed equally as co-first authors to this work.

## Acknowledgments

For financial assisting and technical advice, we thank the project of Shanghai’s Double First-Class University Construction and Development of High-Level Local Universities: Intelligent Medicine Emerging Interdisciplinary Cultivation Project, and Medical Research Data Center of Fudan University.

